# 2020 SARS-CoV-2 diversification in the United States: Establishing a pre-vaccination baseline

**DOI:** 10.1101/2021.06.01.21258185

**Authors:** Adam A. Capoferri, Wei Shao, Jon Spindler, John M. Coffin, Jason W. Rausch, Mary F. Kearney

## Abstract

In 2020, SARS-CoV-2 spread across the United States (U.S.) in three phases distinguished by peaks in the numbers of infections and shifting geographical distribution. We investigated the viral genetic diversity in each phase using sequences publicly available prior to December 15^th^, 2020, when vaccination was initiated in the U.S. In Phase 1 (winter/spring), sequences were already dominated by the D614G Spike mutation and by Phase 3 (fall), genetic diversity of the viral population had tripled and at least 54 new amino acid changes had emerged at frequencies above 5%, several of which were within known antibody epitopes. These findings highlight the need to track the evolution of SARS-CoV-2 variants in the U.S. to ensure continued efficacy of vaccines and antiviral treatments.

**One Sentence Summary:** SARS-CoV-2 genetic diversity in the U.S. increased 3-fold in 2020 and 54 emergent nonsynonymous mutations were detected.

## Main Text

The first reported case of COVID-19 in the U.S. was on January 20^th^, 2020 (*1*). Since that time, through May 2021, there have been more than 32.9 million U.S. cases (20.2% of global) and 585,700 deaths (17.4% of global) (*2*). Several vaccines have been developed, tested, approved, and are now being administered in the U.S. and elsewhere. Although vaccine administration began less than a year after the first reported case in the U.S., expansive global spread of SARS-CoV-2 over this period allowed for the concomitant emergence of variants with greater transmissibility and virulence as well as partial resistance to current preventives and treatments (*3, 4*). Though they share some common genetic features, such ‘Variants of Concern’ (VOCs) appear to have emerged independently in different regions throughout the world, raising the question of whether, and how quickly, variants resistant to induced immunity will evolve in the face of new selection pressures imposed by widespread vaccine administration (*5, 6*).

The spread of SARS-CoV-2 in the U.S. in 2020 occurred in three phases marked by peaks in case numbers, hospitalizations, and deaths (Fig. 1A-B). Phase 1, in the winter and spring of 2020, began with the introduction of SARS-CoV-2 from Europe and Asia (*7*) and was followed by a surge of cases resulting from community spread and mobility (*8*) especially in the Northeast (*9-12*) (Fig. 1C-E). Phase 2 began in early June with accelerated community spread primarily in the South and West after mitigation policies were relaxed (Fig. 1D-E). The start of Phase 3 in the fall of 2020 was marked by a surge of transmission in the Midwest (Fig. 1D-E) followed by a nationwide increase, at or near the end of which time public vaccination was initiated (Fig. S1 & Table S1).

**Fig. 1.**
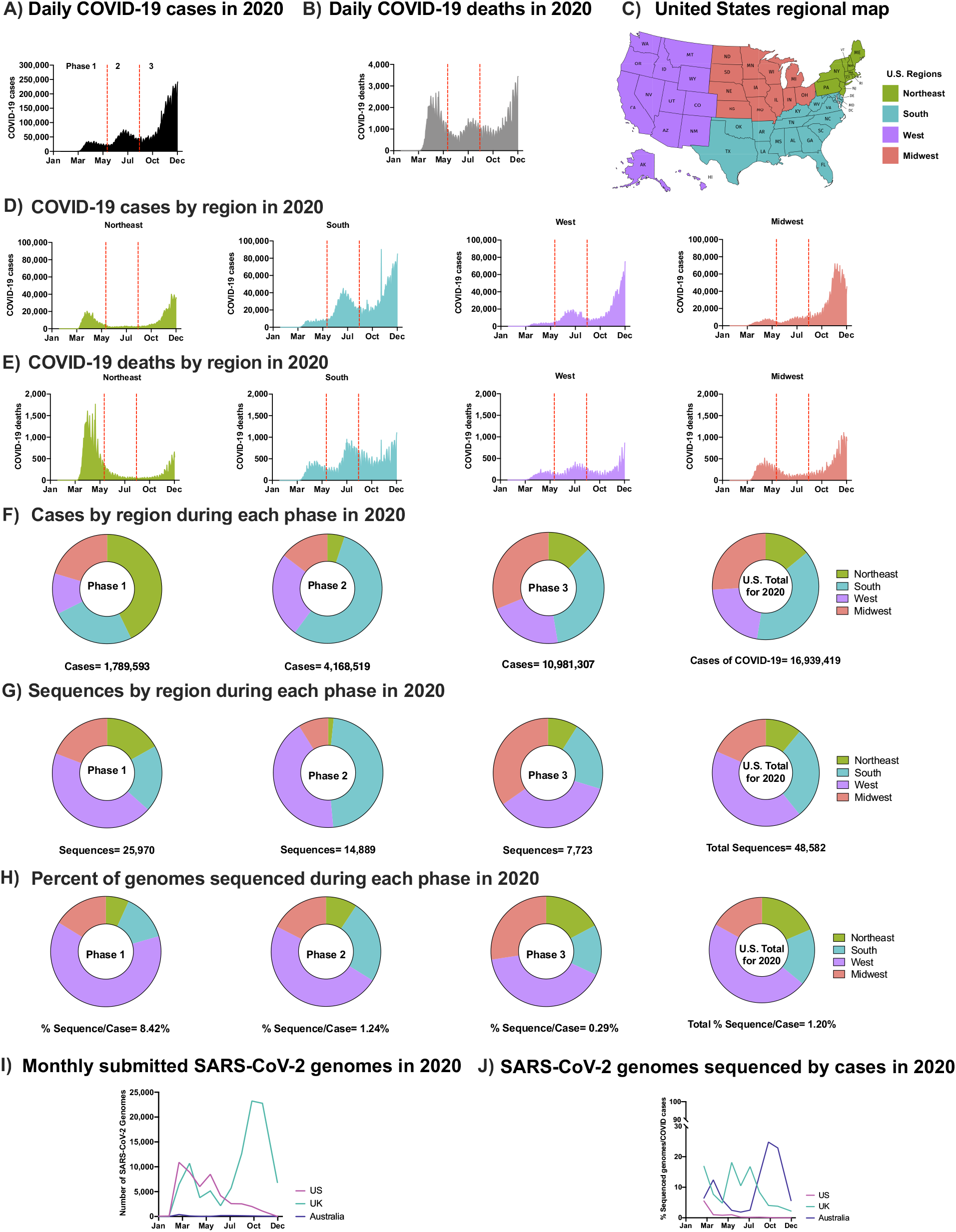
SARS-CoV-2 Epidemic in the U.S. in 2020. (**A**) Daily COVID-19 cases in the U.S. in 2020 (**B**) Daily COVID-19 deaths in the U.S. in 2020 (**C**) U.S. regional map colored by region (**D**) Number of COVID-19 cases in the U.S. in 2020 by region: Northeast, South, West, Midwest, respectively. (**E**) Number of COVID-19 deaths in the U.S. in 2020 by region. (**A-B & D-E**) Separation of Phases is denoted by vertical dotted red lines. Data were smoothed by a moving 3-day average. (**F**) Proportion of COVID-19 cases by region during each phase and the overall contribution to the U.S. total in 2020. (**G**) Proportion of SARS-CoV-2 sequences accessed (submission as of December 15^th^, 2020) by region during each phase and the overall contribution to the U.S. total in 2020 (**H**) The number of sequneces per case were obtained by each region during each phase and the U.S. total in 2020. (**F-H**) Highlights Phase 1, 2, and 3, followed with U.S. total of 2020. (**I**) Total number of sequences submitted to GISAID from the U.K., Australia, and the U.S. by December 15^th^, 2020. (**J**) Submitted SARS-CoV-2 genomes normalized to the number of COVID-19 cases from the U.K., Australia, and the U.S. (see Supplementary Methods).

The regional distribution of COVID-19 cases varied by phase and was not always correlated with the level of viral sequencing in the different regions. For example, although the South region had the greatest overall number of cases (Fig. 1F), the majority of SARS-CoV-2 sequences were obtained from samples collected in the West (Fig. 1G-H). In total, viral sequences were obtained from 1.2% of reported U.S. cases in 2020, compared to 8.1% in the U.K. and 6.2% in Australia (Fig. 1I-J). The aggregate rate of sequencing in the U.S. reflects a decrease from 8.4% in Phase 1 to 0.3% in Phase 3, a difference that can be partly explained by the long intervals between sample collection and sequence deposition in GISAID (median: ∼100 days; Fig. 1H-I). Of note, the rate of sequencing has significantly increased in 2021 compared to 2020.

To characterize the increasing genetic diversity and divergence of SARS-CoV-2 in the U.S. prior to both the introduction of vaccines and detection of the first VOC, high-coverage full-length genome sequences with GISAID submission dates on or before December 15^th^, 2020 were analyzed. All early GISAID-assigned clades of SARS-CoV-2 (G/GH/GR/S/L/V) were identified in the U.S. in Phase 1. However, the G-based clades (G/GH/GR), defined by the D614G mutation in the Spike (S) gene (*13, 14*), accounted for >99% of sequences by Phase 2 (Fig. S2). SARS-CoV-2 variants with the D614G mutation have been shown to be more infectious and exhibit some degree of resistance to certain monoclonal antibodies (*15*), yet they maintain convalescent serum neutralization sensitivity (*16*) and do not appear to worsen clinical outcomes (*17*).

The aggregate average pair-wise distance (APD) among G-based clades increased from 0.02% in Phase 1 to 0.06% in Phase 3, reflective of 2.3-, 3.0-, and 2.8-fold increases for clades G, GH, and GR, respectively (Fig. 2A & Fig. S3). Additionally, the approximate rate of change in APD in the G clades was 1.95-nt/month (clade G), 2.85-nt/month (clade GH), and 2.22-nt/month (clade GR) (Fig. S3A). In contrast, the APD of sequences comprising the VOCs (B.1.1.7, P.1, B.1.351, and B.1.427/429) was between 0.02% and 0.06%, and were within the interquartile range of the diversity measured in the G clades in the U.S. in 2020 despite being introduced towards 2021 (Fig. S3B). The observed increase in genetic diversity of SARS-CoV-2 in the U.S. in 2020 was due to an increase in the number of unique variants comprising clade G (+14%) and clade GR (+17%) (Fig. 2A). However, despite the 3-fold increase in APD, clade GH had an overall decrease in the number of different variants from Phase 1 to Phase 3 (−11%) (Fig. 2A). This finding suggests that the increase in genetic diversity in the GH clade resulted not from an increase in the number of variants but rather from the spread of fewer variants that were more highly divergent. This finding is further supported by observations of the numbers of mutations observed per sequence over time for each clade (Fig. 2B). Specifically, whereas in Phase 1 clades G, GH, and GR averaged 7, 7, and 10 mutations/sequence, respectively, these frequencies increased by 1.7-, 2.4-, and 1.8-fold in Phase 3, another indication of a disproportionate increase in GH clade divergence. For comparison, the VOCs had 26-36 mutations/sequence compared to their clade consensus (Fig. 2B-C). Similarly, panmixia, a metric indicating the degree to which random populations remain unstructured (*i*.*e*., non-divergent) over time (*18*), was calculated for the respective clades. We found there was significant divergence in both the G- and S-based clades from Phase 1 to Phase 2 (*p<10*^*-6*^), demonstrating there was not random unstructured population mixing and that viral evolution was directional (Fig. 2A).

**Fig. 2.**
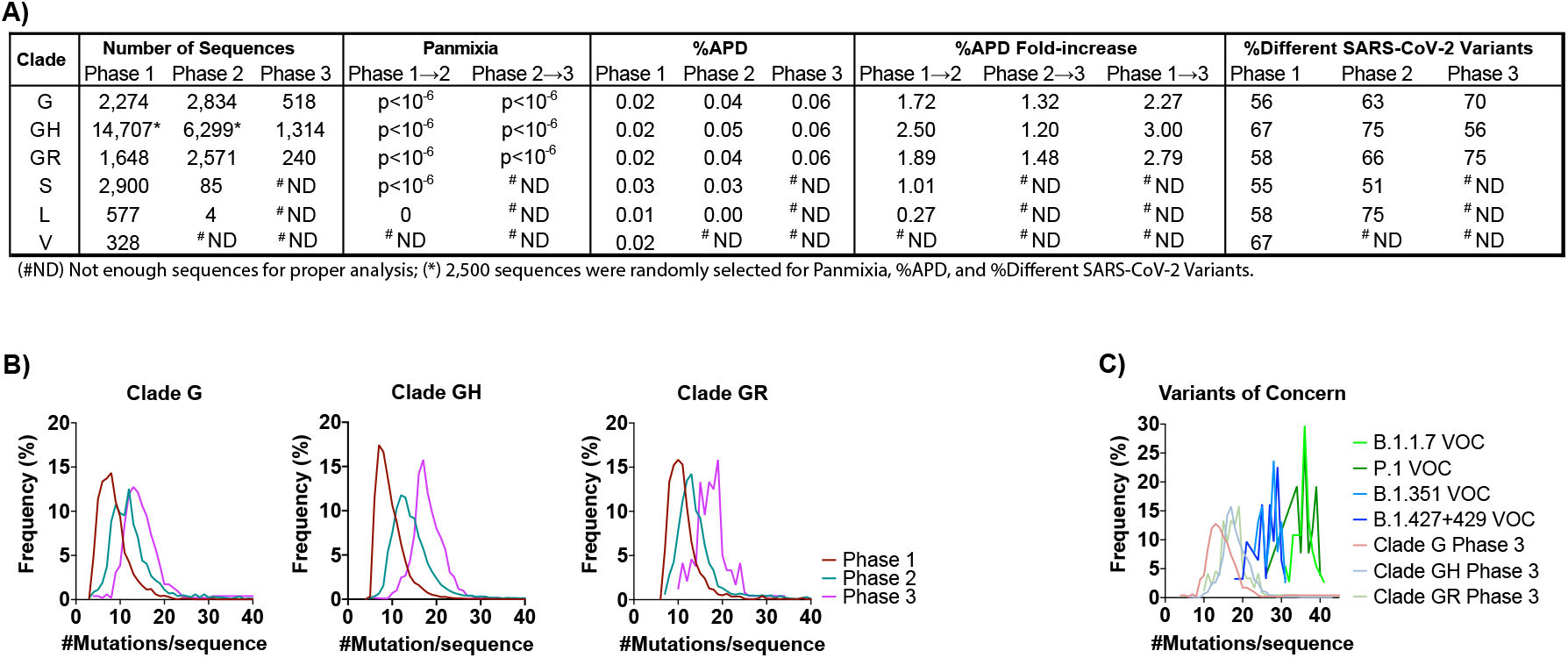
SARS-CoV-2 genetic diversity increased over time. (**A**) The number of sequences obtained from GISAID (https://www.gisaid.org/) in the analysis for each clade by Phase are reported. Genetic divergence was measured by panmixia probability (*18*) (significance cutoff, p<10^−3^) for clade/Phase with >11 sequences for Phase 1→2 and Phase 2→3. Genetic diversity was measured by average pair-wise distance (%APD) and the percent of different SARS-CoV-2 variants. (**B**) The distribution of the frequency for the number of mutations per sequence relative to the Wuhan-Hu-1 isolate was determined for the G-based clades in each Phase. (**C**) Distribution of the frequency for the number of mutations per sequence for the VOCs: B.1.1.7 (20I/501Y.V1, GR, first observed in U.K.), P.1 (20J/501Y.V3, GR, Brazil), B.1.351 (20H/501Y.V2, GH, South Africa), and the collective B.1.427/429 (20C/S:452R, GH, U.S.-California). (^#^ND) Not enough sequences for proper analysis; (*) 2,500 sequences were randomly selected for Panmixia, %APD, and %Different SARS-CoV-2 Variants.

Levels of SARS-CoV-2 sequencing in the U.S. determine both the sensitivity with which we are able to detect emerging variants and our degree of statistical confidence that we have done or can do so. From U.S. viral sequences submitted to GISAID prior to Dec 15, 2020, we were able to detect mutations present in the population at frequencies ≥5% in Phases 1 and 2 and ≥14.5% in Phase 3 with 95% confidence (see Supplementary Methods). Increasing the sensitivity of and confidence in this type of analysis in 2021 will require greater sampling as is now being pursued. For instance, to detect mutations present in each daily sample at 1% or 0.1% frequency with 99% confidence, the viral sequencing capacity in the U.S. needs to increase by ∼7.8-fold (to 460 sequences/day) or ∼78-fold (4,600 sequences/day), respectively (Fig S4), a goal that is now within reach due to increases in funding for SARS-CoV-2 sequencing. For this study, majority-rule consensus sequences of each G clade from Phase 1 were compared to all respective sequences in the subsequent two phases (Table S2-4). Only mutations present in ≥5% of the population were included in the analyses, although mutations of specific interest present at lower frequencies were also noted and are discussed below.

About half of the new mutations that arose in the U.S. and persisted at frequencies ≥5% were nonsynonymous, *i*.*e*., clade G: ORF1a (9 nonsynonymous mutations), ORF1b (4), Spike (1), Nucleocapsid (3); clade GH: ORF1a (12), ORF1b (6), Spike (1), Nucleocapsid (6); and clade GR: ORF1a (5), ORF1b (2), Spike (4), Matrix (1). While some mutations arose and then declined during 2020 (or shifted geographically), most persisted and increased in frequency (Fig. 3A-C). In particular, clade G mutation N^S194L^ and clade GH ORF1a^L3352F^, ORF1b^N1653D;R2613C^ increased more than 40% from Phase 1 to Phase 3. Many new mutations were detected in Phase 3 despite the limitations of extremely shallowing sampling.

**Fig. 3.**
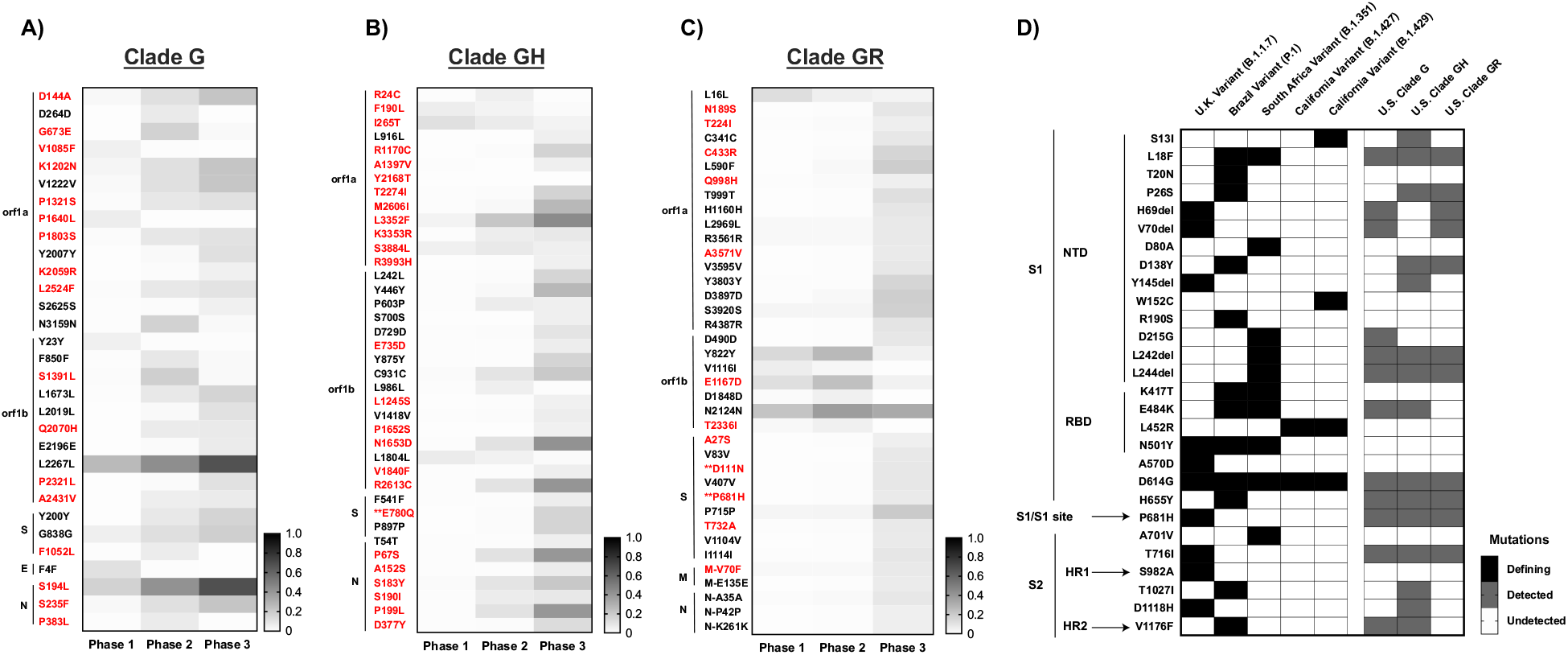
Emerged mutations in SARS-CoV-2 have increased in the U.S. (**A-C**) Non-clade defining mutations of Clades G, GH, and GR. Emerging mutations are shown in which, during at least one Phase in 2020 the frequency exceeded 5%. Sequences were compared to the majority-consensus sequence for each respective clade in Phase 1. Mutation designations reflect the relative amino acid positions in the gene regions. There were no common mutations among the G-based clades. Red text denotes non-synonymous mutations. (**) Mutations that occur in T- or B-cell epitope regions. (**D**) Comparison of non-synonymous mutations and deletions in Spike between VOCs and the U.S. G-based clades across all three phases. The presence or absence of mutations is indicated by shading: VOC-defining mutations (black), mutations detected during at least one phase in the respective G-clade (gray), and undetected in either VOC or U.S. G-clades (white).

Since all current vaccines were designed to potentiate cellular and humoral immune memory responses against the SARS-CoV-2 Spike protein (*19*), the emergence of mutations in the viral gene encoding Spike warrant special attention. In accordance with this elevated level of importance, VOCs are defined primarily, though not exclusively, by their associated Spike mutations. Critical investigations have been and are being conducted to determine how emergent Spike mutations affect vaccine efficacy (*20, 21*), as well as effectiveness of immunotherapeutics (*i*.*e*., monoclonal antibodies and convalescent plasma) (*3, 21*), and viral transmission and pathogenesis (*13, 17, 22*). Selection of Spike mutations among recipients of convalescent plasma is particularly concerning (*23-25*), since the partial protection conferred by such treatments is likely conducive to immune escape, and may have contributed to the emergence of the B.1.1.7, B.1.351, P.1, and B.1.427/429 VOC (*26*).

Although the VOCs emerged later in 2020, our analysis shows that many of their defining mutations were present individually in the U.S. at low frequency (<1% of total sequences) as early as Phase 1 (Fig. 3D). However, with the exception of S^P681H^, the prevalence of these mutations never reached 5% in any of the 3 Phases, and they were therefore excluded from our primary analysis. Mutations defining the B.1.427/429 VOC (S^S13I;W152C;L452R^) which originated in the U.S., were only detected in a very small fraction of sequences in Dec 2020, possibily due to the lag between sample collection and sequence deposition and/or shallow Phase 3 sampling. The S^E484K^ mutation has been identified in multiple VOCs and is particularly concerning, having also been reported to contribute to immune evasion (*3*). At this time, however, immunity induced by mRNA-based vaccines appears to remain at least partially effective against S^E484K^ – containing VOCs (*27-30*).

Although the replication fidelity of SARS-CoV-2 is quite high relative to other RNA viruses (*5, 31*), its genetic stability is countered by the expansive spread of the virus both in the U.S. and globally. On balance, evolution of this virus might be best characterized as slow but inexorable, driven largely by genetic drift but also influenced by selective pressures such as relative infectivity, relative transmissibility, and perhaps even immune evasion. Conversely, the relatively recent and suddenly emergent VOCs are characterized by a significantly higher degree of genetic divergence that is rapidly acquired and clearly confers a replicative advantage. These variants were most likely the product of isolated cases in which the evolutionary environments differ substantially from the norm; *e*.*g*., from chronically infected immunosuppressed individuals including those who received treatment with monoclonal antibodies or convalescent serum (*23, 25, 32*).

Regardless of the evolutionary pathways by which they emerge, the emergence of mutations in both the U.S. and global viral populations threatens the continued efficacy of current treatments and vaccines and will do so increasingly as the virus continues to spread and evolve. In response, scientists worldwide are coming together to increase timely SARS-CoV-2 sequencing and analysis to help guide decisions on current and future health policies. Unfortunately, in 2020, efforts to slow the spread of the virus within the U.S. was hampered by low adherence to recommended preventative measures (*e*.*g*., mask wearing, hand washing, social distancing) as well as inconsistent and even conflicting messaging regarding these measures. These failures have indirectly accelerated viral divergence, increased genetic diversity, and resulted in the accumulation of nonsynonymous mutations, many of which are within epitopes associated with resistance to neutralizing antibodies that may ultimately contribute to immune evasion.

As more people are vaccinated, selective pressure for immune-resistant variants will increase concomitantly. As such, monitoring individuals that become infected with SARS-CoV-2 after vaccination, and those with prolonged infection (*e*.*g*., immunocompromised individuals) is essential to understanding the capacity of SARS-CoV-2 for escape from vaccine-induced immunity. While our study focused on the U.S., it is important to recognize that the emergence of potential new VOCs is an ongoing and global concern. Therefore, we must maximize our vigilance for detecting and analyzing new variants, and just as importantly, continue to encourage measures to prevent their spread.

## Data Availability

All data is available in the main text or the supplementary materials. In addition, can be accessed on GISAID.org.

## Acknowledgments

We gratefully acknowledge the Authors, the Originating and Submitting Laboratories for their sequence and metadata shared through GISAID, on which this research is based. We would also like to thank the COVID Tracking Project.

## Author Contribution

Conception and design: AAC, JMC, JWR, and MFK. Collection and assembly of data: AAC, WS, JS. Analysis and interpretation of the data: AAC, WS, JMC, JWR, and MFK. Drafting of the article: AAC, JMC, JWR, and MFK. All authors wrote and approved the final article.

## Disclaimer

The content of this publication does not necessarily reflect the views or policies of the Department of Health and Human Services of the Unites States government.

## Data and materials availability

All data is available in the main text or the supplementary materials.

## Supplementary Materials

### Materials and Methods

#### COVID-19 Cases and Deaths

Data for U.S. COVID-19 cases and deaths were extracted from https://covidtracking.com/data from 01-January-2020 through 15-December-2020 (accessed 18-December-2020). U.S. regions were assigned based on the four Census Regions of the United States (U.S. Census Bureau) (*32*). The pandemic in the U.S. was divided into three phases based on COVID-19 case peaks where the derivative of the trough was approximately zero: Phase 1 of winter-spring (01-January-2020—31-Mary-2020), Phase 2 of summer (01-June-2020—31-August-2020), and Phase 3 of Fall (01-September-2020—15-December-2020). The estimated 2019 population for each U.S. State was accessed by the U.S. Census Bureau (Table S1) in order to normalize the incidence of COVID-19 cases and deaths in the sub-regional areas of the U.S. GraphPad Prism V.8.4.3 was used to visualize the data.

#### SARS-COV-2 Sequences

A total of 36,299 full-length (29,782 bp), high-coverage SARS-CoV-2 genomes from humans in the U.S. with infections between 20-January-2020 through 15-December-2020 were obtained from gisaid.org (accessed 18-December-2020) for sequence analysis. The numbers of sequences obtained were: Phase 1 (22,434), Phase 2 (11,893), and Phase 3 (2,072). Any nucleotide position with an ‘N’ was replaced with a gap. SARS-CoV-2 Clade O, Clade GV, Cruise-ship, and other U.S. territory sequences were excluded in the analysis due to the small representation. Variants of Concern (VOC or VOCs) from the United Kingdom (U.K.), South Africa, Brazil, and California were accessed from gisaid.org on 01 April 2021 (see also “Analysis of Variants of Concern” section of methods). The VOCs were sampled in the U.S. between 01 November 2020 through 31 March 2021. Fifty VOCs sequences were randomly selected for analyses and can be found in the **Supplemental File**. Sequences that contained ‘Ns’ were excluded. There was a total of 37 sequences (B.1.1.7 VOC), 38 sequences (B.1.351 VOC), 26 sequences (P.1 VOC), and 31 sequences (B.1.427/429 VOC) for the final dataset. Gap-stripped alignments were generated using the FFT-NS-1 200PAM/k=2 algorithm of MAFFT v7.450 (*33, 34*). Additional analyses of SARS-CoV-2 sequences were done using Geneious Prime® 2020.2.4 (*35*) and an in-house generated pipeline available at https://github.com/Wei-Shao/COV2-Analysis.

#### Distribution and genetic diversity of SARS-COV-2 in the U.S. in 2020

U.S. SARS-COV-2 genomes from each phase were separated by region and clade for each month in 2020. Monthly clade datasets with less than 10 sequences were excluded. To estimate the number of COVID-19 cases within each GISAID clade, the number of sequences was multiplied by the total monthly new COVID-19 cases. RStudio v1.3 (*36*) and GraphPad Prism V.8.4.3 were used to generate the figures.

The rate of SARS-COV-2 sequencing in the U.S. in 2020 was compared to the rate in the U.K. and in Australia. Sequence data for the U.K. and Australia was obtained under the same selection criteria as for the U.S. The number of cases were accessed on the same days using https://coronavirus.data.gov.uk/ and the National Notifiable Diseases Surveillance System (http://www9.health.gov.au/cda/source/rpt_3.cfm). The rate was determined from the number of sequences obtained monthly and the number of monthly cases of COVID-19 using in-house generated bioinformatic pipelines available at https://github.com/aacapoferri/COV2.

#### Genetic characterization and mutation analyses

Using in-house bioinformatic pipelines available at https://github.com/aacapoferri/COV2, SARS-COV-2 mutation frequencies were determined for each clade/Phase compared to either the majority-rule Phase 1 consensus sequence described next, or the Wuhan-Hu-1 reference genome (GenBank accession, NC_045512.2) or the VOCs (*37*). To exclude clade-defining mutations and amplification/sequencing errors, several steps were taken. First, majority-rule consensus sequences for each clade were generated from all genomes in Phase 1 and used as a reference in downstream analyses. This approach allowed majority clade-associated mutations to be omitted for the detection of new mutations only. Second, a threshold ≥5% frequency was used to eliminate mutations that were rarely detected and, therefore, could be PCR, sequencing errors, or real but not indicative of an emerging variant (see also “Number of sequences to detect mutations” section of methods). Mutation frequencies were plotted and annotated using the “Mutation frequency for SARS.R” script (an example provided on https://github.com/aacapoferri/COV2). Mutations that were above the ≥5% threshold were noted for each G-based clade and phase. The heatmaps for clades G, GH, and GR were generated to visualize the persistence and emergence of mutations present at frequencies ≥5% using GraphPad Prism V.8.4.3.

All sequences for each clade and Phase were included in each analysis with the exception of clade GH, where the number of sequences was too high for measurements of genetic diversity and divergence and, therefore, 2,500 sequences were randomly subsampled for those analyses. Mutation distributions were determined by assessing the number of mutations per sequence for each clade during each Phase. Statistical shifts in population structure (divergence) were determined using a test for panmixia with a statistical cut-off at *p<10*^*-3*^ (*38*). Population genetic diversity was calculated as average pair-wise distance (APD) in MEGAX for each clade/Phase (*39*). These calculations were repeated with all clades during each month with at least 10 sequences or randomized sub-sampling of 50 sequences in triplicate to determine the APD. In cases where a particular clade had less than 10 sequences in a given month, they were excluded from analysis. Identical SARS-COV-2 genomes were collapsed to determine the number of different variants in the dataset. A simple linear regression was determined for clades G, GH, and GR in GraphPad Prism V.8.4.3 with the linear equation and goodness of fit R^2^ reported. The slope was understood as the rate of change in %APD/month. The length of the SARS-CoV-2 genome is ∼30,000 base pair, which when multiplied by the slope, gave an approximate number of nucleotide changes/month for a given G-based clade.

To examine the number of mutations per sequence in the G-based clades during each phase, the distribution of the number of mutations relative to the Wuhan-Hu-1 reference genome was plotted using in-house pipelines available at https://github.com/Wei-Shao/COV2-Analysis by Hamming distances.

#### Analysis of Variants of Concern (VOCs)

The four VOCs used in this study included the Pango lineage B.1.1.7 (Nexstrain 20I/501Y.V1, GISAID clade GR, originally isolated in the U.K.), P.1 (20J/501Y.V3, GR, Brazil), B.1.351 (20H/501Y.V2, GH, South Africa), and the B.1.427+429 (20C/S:452R, GH, California) sampled at locations in the U.S. After sequence sample processing, the number of mutations per sequence was determined for each VOC. The collection sampling on GISAID was set to any submitted sequences that spanned 5 months (November 2020-March 2021). We wanted to ensure that all VOCs circulating in the U.S. would be captured. The individual VOC sequences collected for the final dataset in the U.S. were identified for 37 sequences of B.1.1.7 (February-March 2021), 26 sequences of P.1 (January-March 2021), 38 sequences of B.1.351 (January-March 2021), and 31 sequences of B.1.427+429 (December 2020 and January-February 2021).

The distribution of the number of mutations per sequence was compared to each respective derivative GISAID clade distribution during Phase 3, which was closest to the emergence of the VOC. The overall APD for each VOC dataset was calculated and compared to the genetic diversity of the G-based clades in the U.S. during 2020.

Due to the interest in the Spike protein of SARS-CoV-2 for vaccine and therapeutic strategies, VOC defining mutations in the S gene were specifically explored and were compared to sequences obtained from each Phase of infections in 2020. For this analysis mutations at frequencies less than 5% were included.

#### Potential effect of mutations

Majority-rule consensus sequences were generated for the G-based clades at each phase and were aligned to the Wuhan-Hu-1 reference genome. Previously mapped T-cell and B-cell epitopes from Spike and Nucleocapsid were annotated on the reference genome and included in Table S5 Nonsynonymous mutations observed in the G-based clades that differed from the reference in either T-cell or B-cell epitopes were noted.

#### Viral genetic surveillance resources

Several databases were used to compare and contrast global trends and our U.S.-based specific analysis. These included: PANGO lineages (https://cov-lineages.org/), NextStrain (https://nextstrain.org/sars-cov-2/), Global Initiative on Sharing Avian Influenza Data (https://www.gisaid.org/), Outbreak.info (https://outbreak.info/situation-reports), Coronavirus Resource Center (https://coronavirus.jhu.edu/map.html), Observable (https://observablehq.com/@spond/linkage-disequilibirum-in-sars-cov-2), Virological forum (https://virological.org/), Los Alamos National Laboratory (https://cov.lanl.gov/content/index), and the U.S. Centers for Disease Control and Prevention (https://www.cdc.gov/coronavirus/2019-ncov/cases-updates/variant-surveillance/variant-info.html).

## Supplemental Figures and Tables

**Fig. S1.**
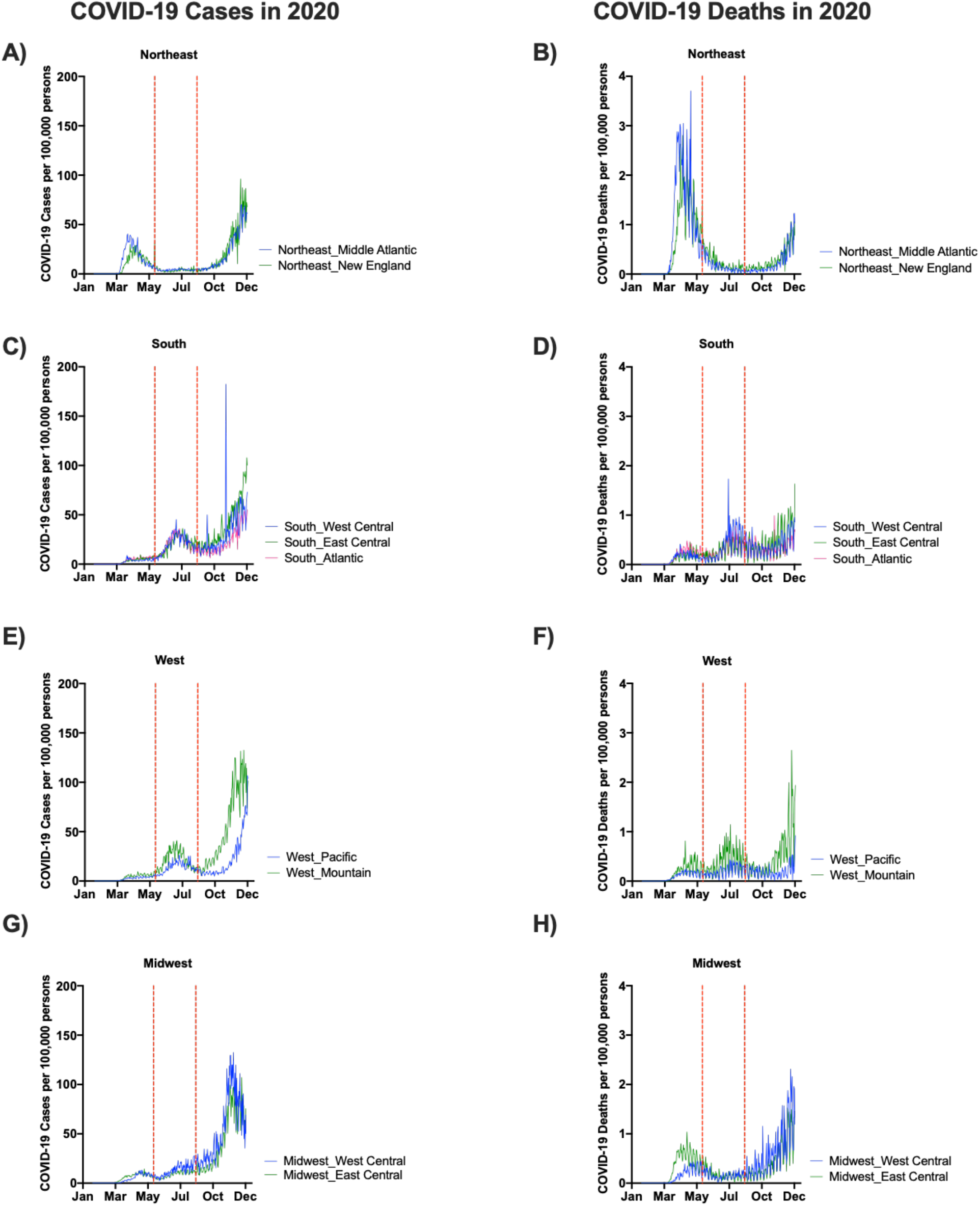
SARS-CoV-2 epidemic at the divisional level in the U.S. (**A, C, E, G**) Number of daily COVID-19 Cases in 2020 per 100,000 persons normalized to the 2019 estimated population in each sub-region. (**B, D, F, H**) Number of daily COVID-19 Deaths in 2020 per 100,000 persons normalized to the 2019 estimated population in each sub-region. (**A-B**) Northeast divisions of Middle Atlantic and New England. (**C-D**) Southern divisions of West Central, East Central, and Atlantic. (**E-F**) Western divisions of the Pacific and Mountain. (**G-H**) Midwestern divisions of West Central and East Central. Each division is colored respectively with dotted red lines indicating the separation of Phases. Population and date of first COVID-19 case in each sub-region is report in **Table S1**.

**Fig. S2.**
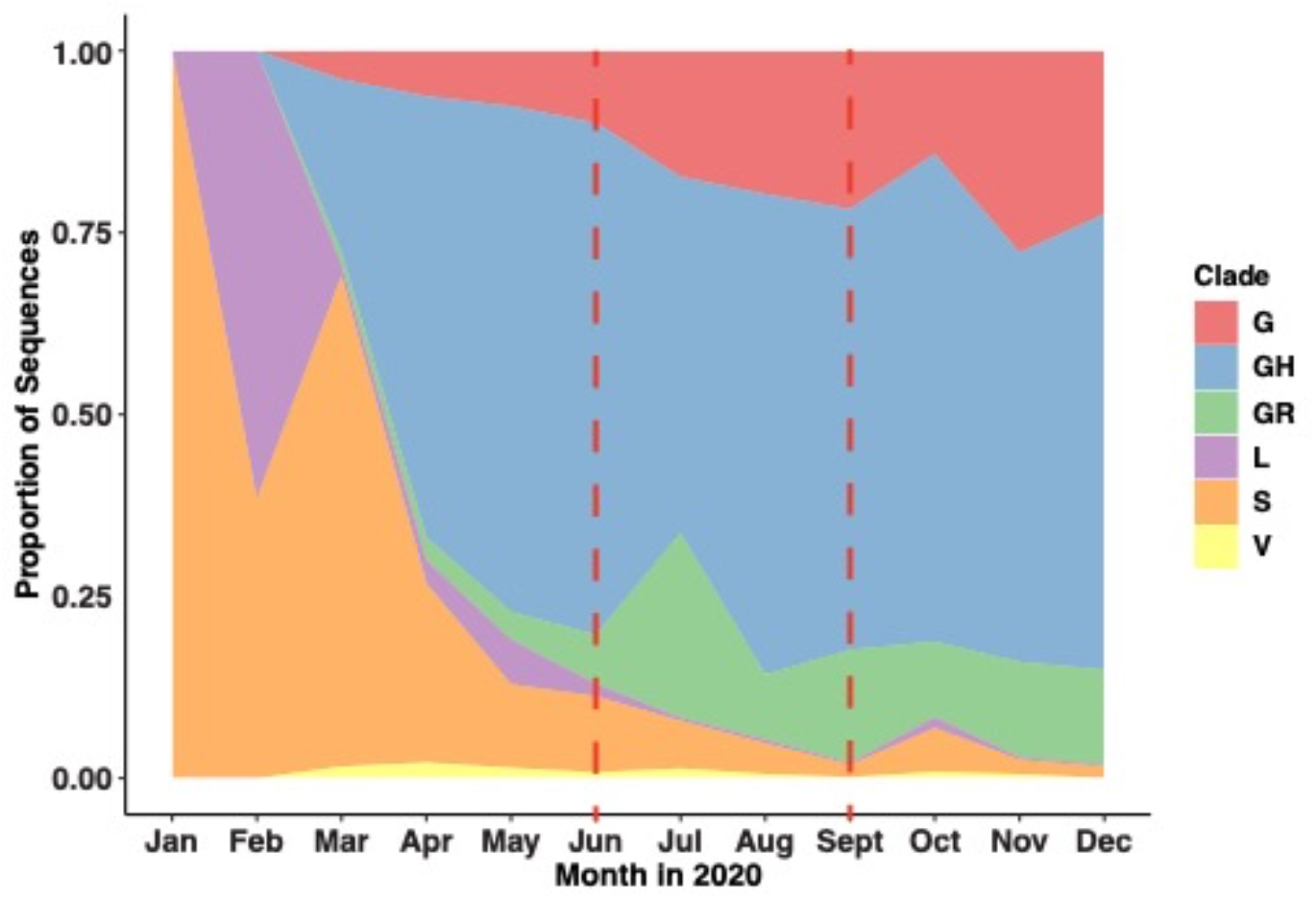
Proportion of SARS-CoV-2 genomes in each GISAID clade. The proportion of sequences observed during each month in 2020 based on the GISAID assigned Clades was calculated. The dotted red lines indicate the separation of Phases 1, 2, and 3. Data were accessed 18-December-2020 whereby any sequences submitted or collected by 15-December-2020 were considered. Clades G, GH, and GR represented the majority of all sequences by Phase 3.

**Fig. S3.**
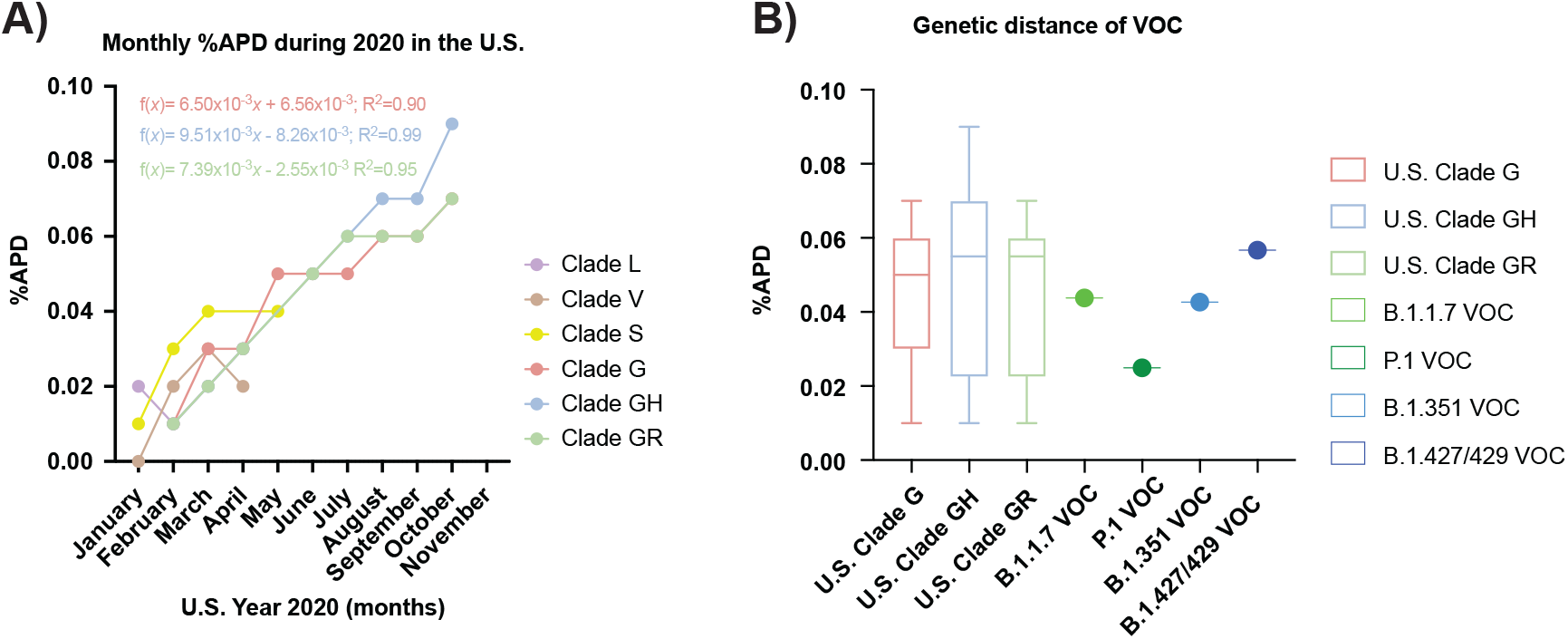
Genetic distance over time in 2020 for GISAID clades and Variants of Concern. (**A**) Average pairwise distance (APD) was calculated per month for sequences within each clade. Time points with less than 10 sequeunces were excluded. Three randomized subsamples of 50 sequences each were analyzed and the APD was calculated to ensure consistency between each subsampling. If there were 11-50 sequences for a clade at a given month, no subsampling was performed. A standard linear regression was run for clades G, GH, and GR. The rate of change in %APD over time is noted in the figure with the goodness of fit (R^2^) reported. The %APD was plotted according to each respective month. The rate of change was 6.50×10^−3^ APD/month (clade G), 9.51×10^−3^ APD/month (clade GH), and 7.39×10^−3^ APD/month (clade GR); whch corresponded to 1.95 nt/month (clade G), 2.85 nt/month (clade GH), and 2.22 nt/month (clade GR) based on the SARS-CoV-2 genome of ∼30,000 bp. (**B**) Comparison of the %APD between the VOCs and the G-based Clades. A box and whister plot of the %APD of the G-based clades in the U.S. during 2020 is shown. The median (solid line) is marked within the interquartile range, with the whiskers as the minimum and maximum. The VOC sequences sampled in the U.S. were accessed through GISAID.org with the collection date between 1 November 2020 to 31 March 2021. Total APD was calculated for the VOCs.

**Fig. S4.**
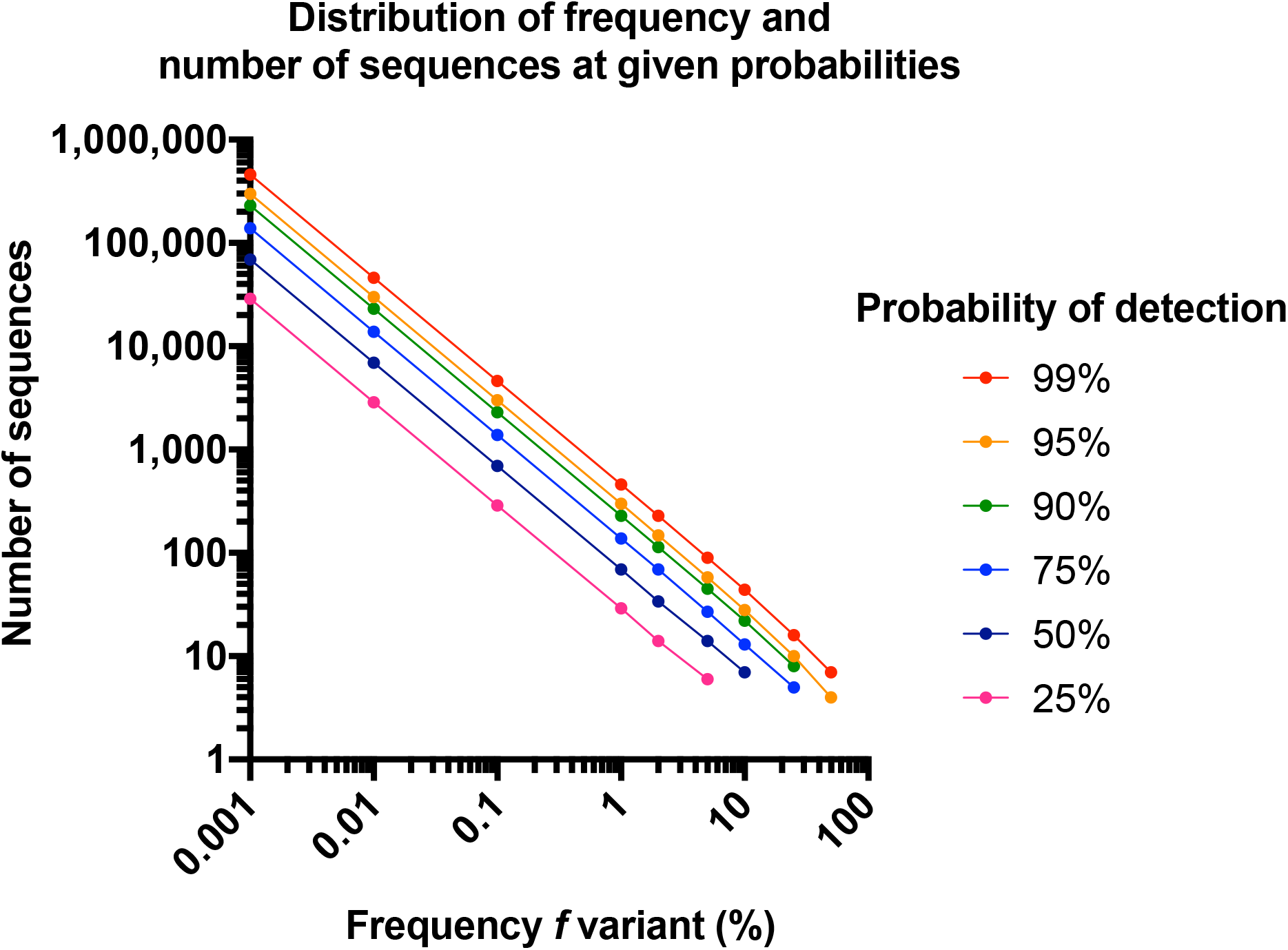
Determining number of sequences of sampling at a given probability to detect variant frequency. To determine the number of sequences required to detect at a certain mutation frequency with a given probability, follows the quotient: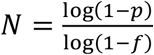. Where (*N*) is the number of sequences of any given sample that is required to be sampled to determine (*f*), the frequency the mutant detected, at a given (*p*), the probability of detecting the mutation at the provided frequency.

**Fig. S5.**
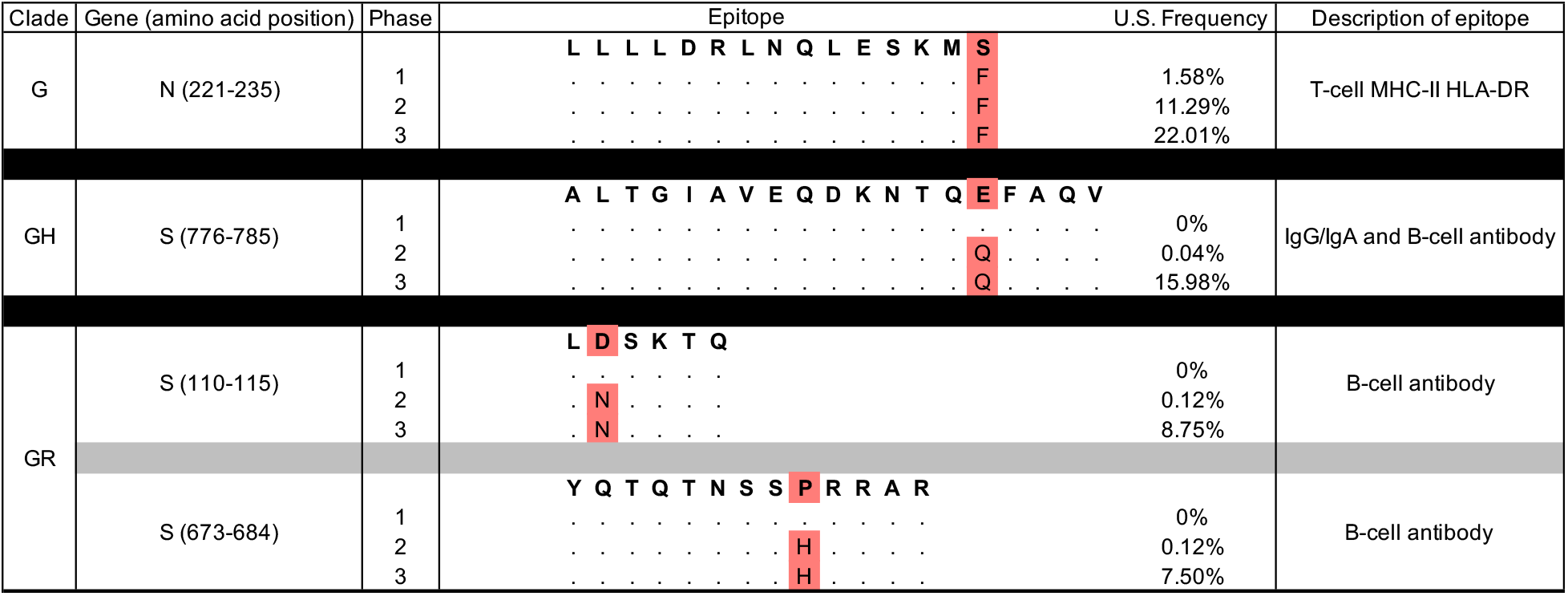
Mutations in G clades that are located in MHC-II HLA-DR T-cell and B-cell epitopes. Nucleocapsid MHC-I/II T-cell and Spike B-cell epitopes with detected mutations are shown. Nonsynonymous mutations were detected in Clades GH and GR starting in Phase 2 through Phase 3 (highlighted). The amino acid position of the epitope is denoted next to N (Nucleocapsid) and S (Spike). The U.S. frequency of detection is noted for each Phase. Epitopes examined are defined by **Table S5**.

**Table S1.**
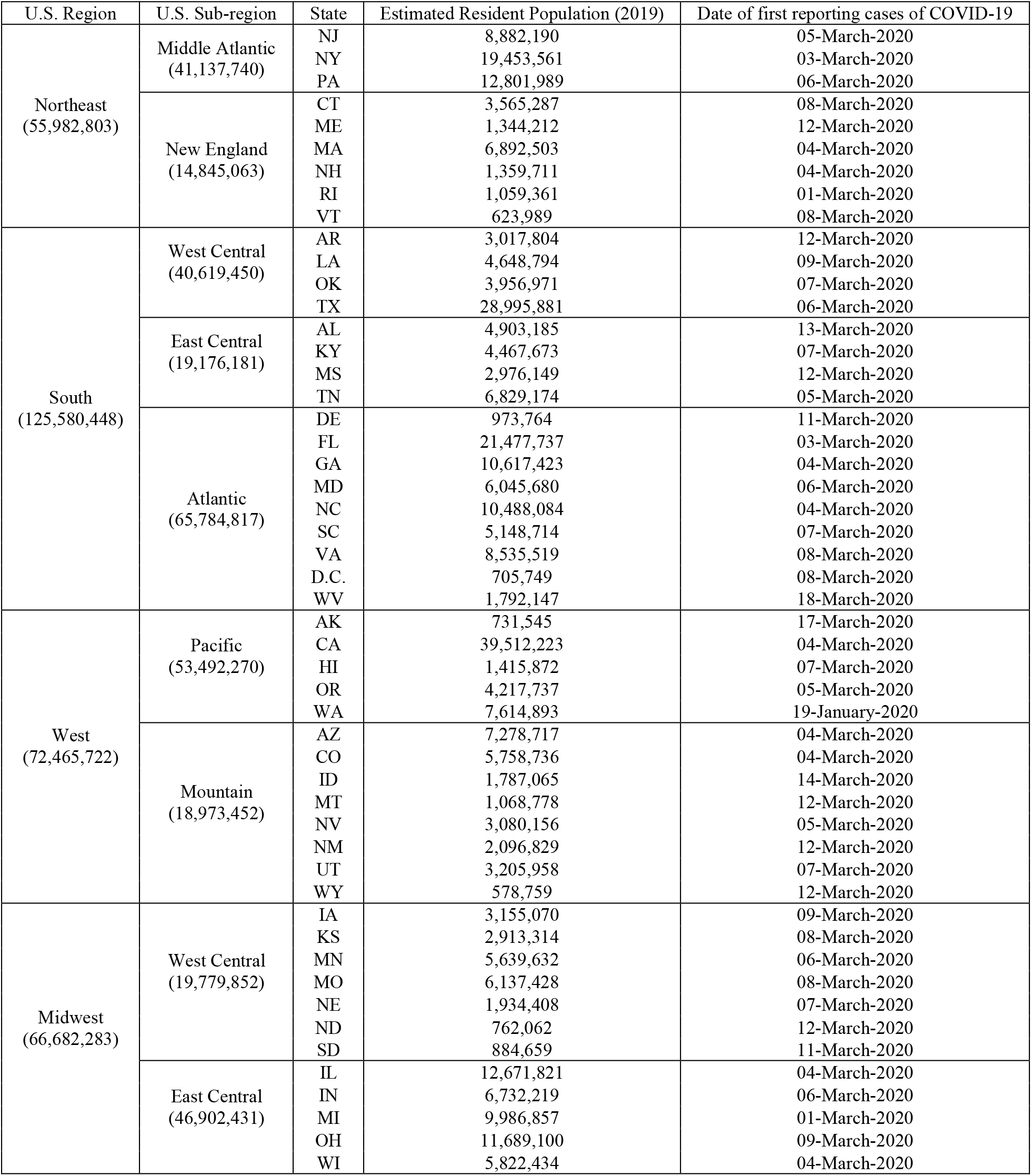
Demographics and Regional Division based on the U.S. Census Bureau. The 2019 estimated residential population for each State is reported. The first reported confirmed plus probable cases of COVID-19 are included (https://covidtracking.com/). The U.S. is generally divided into the Northeast, South, West, and Midwest. Where each major region can be divided into sub-regions. These estimated populations were then used to normalize the incidence of COVID-19 cases and deaths in the U.S. at the sub-regional level.

**Table S2.** Non-associated Clade G mutations that either persisted or emerged during 2020. Data were extracted where, during at least one Phase period, the frequency of a particular mutation was ≥5% compared to the clade Phase 1 majority consensus. The mutation specifies the nucleotide change as well as the amino acid change with coordinates of the gene and its product. Non-synonymous mutations are shown in red.

| Gene | Mutation | Frequency (%) |  |  | Net Change (%) |
| --- | --- | --- | --- | --- | --- |
|  |  | Phase 1 | Phase 2 | Phase 3 | Phase 1→3 |
| orf1a | a431c(D144A) | 1.72 | 11.29 | 21.81 | +20.09 |
|  | c792t(D264D) | 0 | 7.59 | 0 | 0 |
|  | g2017a(G673E) | 0 | 16.83 | 1.54 | +1.54 |
|  | g3253t(V1085F) | 5.1 | 0.07 | 0 | -5.10 |
|  | g3606t(K1202N) | 3.21 | 11.29 | 21.81 | +18.60 |
|  | t3666c(V1222V) | 1.54 | 11.04 | 21.62 | +20.08 |
|  | c3961t(P1321S) | 0.40 | 8.68 | 11 | +10.60 |
|  | c4919t(P1640L) | 6.02 | 0 | 0 | -6.02 |
|  | c5407t(P1803S) | 0.53 | 8.82 | 10.62 | +10.09 |
|  | c6020t(Y2007Y) | 0 | 1.24 | 11.20 | +11.20 |
|  | a6176g(K2059R) | 0 | 0.49 | 5.02 | +5.02 |
|  | a7572c(L2524F) | 0.40 | 8.57 | 10.23 | +9.83 |
|  | c7875t(S2625S) | 0.09 | 0.49 | 5.02 | +4.93 |
|  | t9477c(N3159N) | 0.13 | 17.29 | 1.54 | +1.41 |
| orf1b | c69t(Y23Y) | 5.01 | 0.21 | 0.39 | -4.62 |
|  | c2550t(F850F) | 0.31 | 9.03 | 1.35 | +1.04 |
|  | c4172t(S1391L) | 0.40 | 18.38 | 1.54 | +1.14 |
|  | c5019t(L1673L) | 0.04 | 8.54 | 15.83 | +15.79 |
|  | c6057t(L2019L) | 0 | 1.2 | 10.81 | +10.81 |
|  | g6210t(Q2070H) | 0 | 6.99 | 7.53 | +7.53 |
|  | a6588g(E2196E) | 0 | 1.09 | 6.76 | +6.76 |
|  | a6801g(L2267L) | 25.99 | 43.9 | 67.37 | +41.38 |
|  | c6962t(P2321L) | 0 | 1.13 | 10.81 | +10.81 |
|  | c7292t(A2431V) | 0 | 5.75 | 7.14 | +7.14 |
| S | t600c(Y200Y) | 0 | 8.57 | 15.83 | +15.83 |
|  | t2514c(G838G) | 5.5 | 12.14 | 18.15 | +12.65 |
|  | c3156a(F1052L) | 0.09 | 6.67 | 0 | -0.09 |
| E | c12t(F4F) | 10.82 | 0.18 | 0 | -10.82 |
| N | c581t(S194L) | 17.19 | 41.78 | 68.34 | +51.15 |
|  | c704t(S235F) | 1.58 | 11.29 | 22.01 | +20.43 |
|  | c1148t(P383L) | 0.13 | 6.81 | 0.19 | +0.06 |

**Table S3.** Non-associated Clade GH mutations that either persisted or emerged during 2020. Data were extracted where, during at least one Phase period, the frequency of a particular mutation was ≥5% compared to the clade Phase 1 majority consensus. The mutation specifies the nucleotide change as well as the amino acid change with coordinates of the gene and its product. Non-synonymous mutations are shown in red,

| Gene | Mutation | Frequency (%) |  |  | Net Change (%) |
| --- | --- | --- | --- | --- | --- |
| | | Phase 1 | Phase 2 | Phase 3 | Phase 1 → 3 |
| orf1a | c70t(R24C) | 0.05 | 5.35 | 0.23 | +0.18 |
|  | t568c(F190L) | 6.03 | 3.98 | 0.91 | -5.12 |
|  | t794c(I265T) | 11.29 | 7.40 | 4.19 | -7.1 |
|  | g2748a(L916L) | 0 | 0 | 5.18 | +5.18 |
|  | c3508t(R1170C) | 0 | 0.65 | 16.36 | +16.36 |
|  | c4190t(A1397V) | 0.03 | 0 | 5.18 | +5.15 |
|  | c6821t(T2274I) | 0.02 | 0.44 | 16.21 | +16.19 |
|  | g7818a(M2606I) | 0.03 | 2.16 | 27.09 | +27.06 |
|  | c10054t(L3352F) | 3.98 | 21.53 | 44.67 | +40.69 |
|  | a10058g(K3353R) | 0.19 | 6.10 | 8.60 | +8.41 |
|  | c11651t(S3884L) | 4.73 | 8.26 | 5.18 | +0.45 |
|  | g11978a(R3993H) | 0 | 0.02 | 5.10 | +5.1 |
| orf1b | t724c(L242L) | 0.01 | 0.67 | 15.60 | +15.59 |
|  | c1338t(Y446Y) | 0.05 | 2.19 | 27.55 | +27.5 |
|  | t1809a(P603P) | 0.01 | 6.37 | 5.25 | +5.24 |
|  | t2100c(S700S) | 0 | 0 | 5.18 | +5.18 |
|  | c2187t(D729D) | 0.07 | 0.62 | 9.59 | +9.52 |
|  | g2205t(E735D) | 0.01 | 0.05 | 5.94 | +5.93 |
|  | c2625t(Y875Y) | 0.03 | 0.11 | 16.06 | +16.03 |
|  | c2793t(C931C) | 1.04 | 9.56 | 20.47 | +19.43 |
|  | t2958g(L986L) | 0.01 | 5.27 | 0.23 | +0.22 |
|  | t3734c(L1245S) | 0 | 0.02 | 5.02 | +5.02 |
|  | g4254t(V1418V) | 0.03 | 0.40 | 5.94 | +5.91 |
|  | c4954t(P1652S) | 0 | 0 | 5.02 | +5.02 |
|  | a4957g(N1653D) | 0.04 | 10.57 | 42.09 | +42.05 |
|  | c5410t(L1804L) | 6.78 | 4.38 | 0.99 | -5.79 |
|  | g5518t(V1840F) | 0.01 | 0.37 | 5.78 | +5.77 |
|  | c7837t(R2613C) | 0.06 | 10.02 | 40.64 | +40.58 |
| S | c1623t(F541F) | 0.01 | 0.13 | 6.01 | +6.00 |
|  | g2338c(E780Q) | 0 | 0.41 | 15.98 | +15.98 |
|  | a2691t(P897P) | 0 | 0.64 | 16.06 | +16.06 |
| N | c162t(T54T) | 0.01 | 0.06 | 5.18 | +5.17 |
|  | c199t(P67S) | 0.03 | 10.46 | 39.88 | +39.85 |
|  | g454t(A152S) | 0.01 | 0.08 | 6.32 | +6.31 |
|  | c548a(S183Y) | 1.03 | 9.53 | 20.70 | +19.67 |
|  | g569t(S190I) | 0.01 | 6.32 | 8.37 | +8.36 |
|  | c596t(P199L) | 0.04 | 10.26 | 39.80 | +39.76 |
|  | g1129t(D377Y) | 0.14 | 0.60 | 13.17 | +13.03 |

**Table S4.**
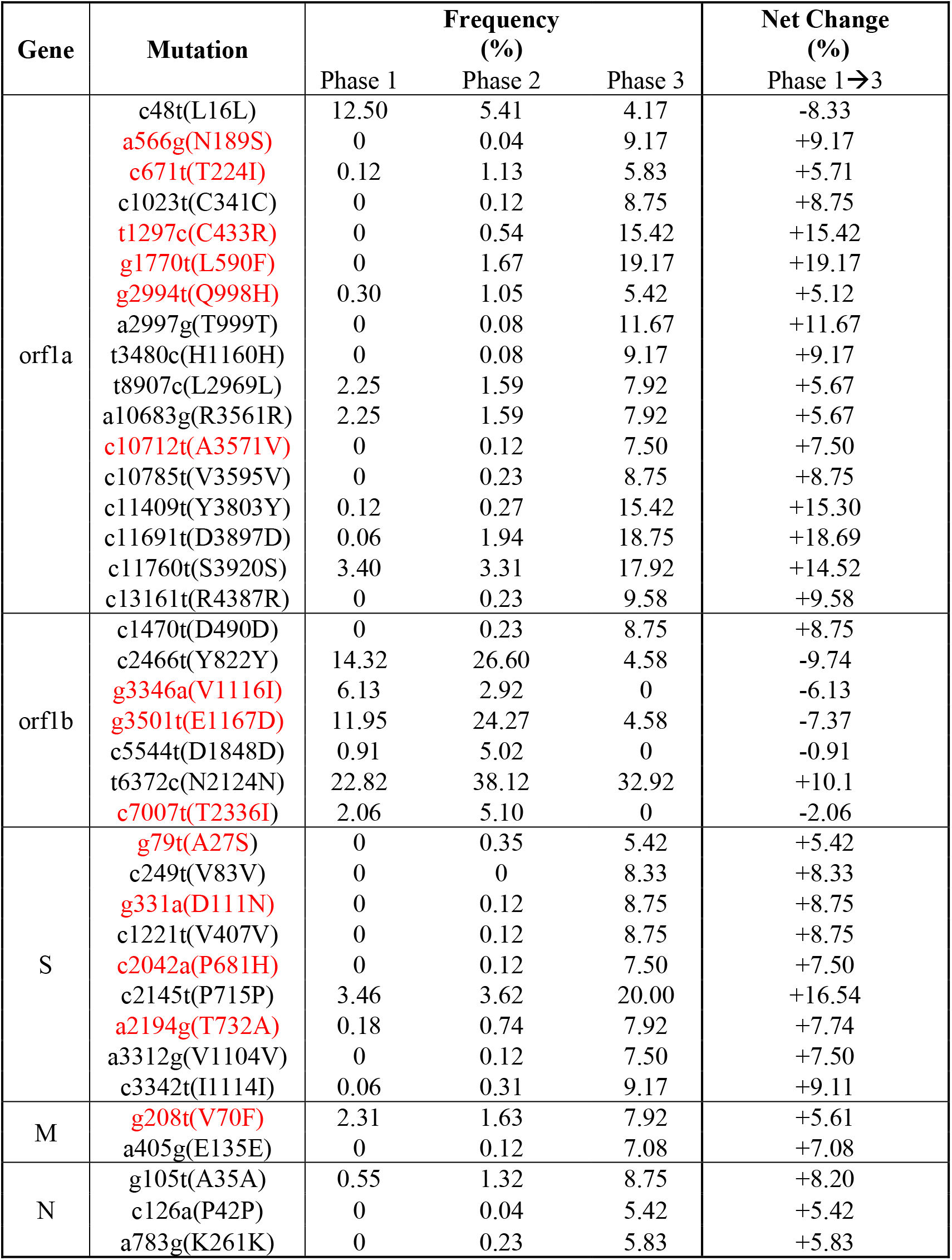
Non-associated Clade GR mutations that either persisted or emerged during 2020. Data were extracted where, during at least one Phase period, the frequency of a particular mutation was ≥5% compared to the clade Phase 1 majority consensus. The mutation specifies the nucleotide change as well as the amino acid change with coordinates of the gene and its product. Non-synonymous mutations are shown in red.

**Table S5.**
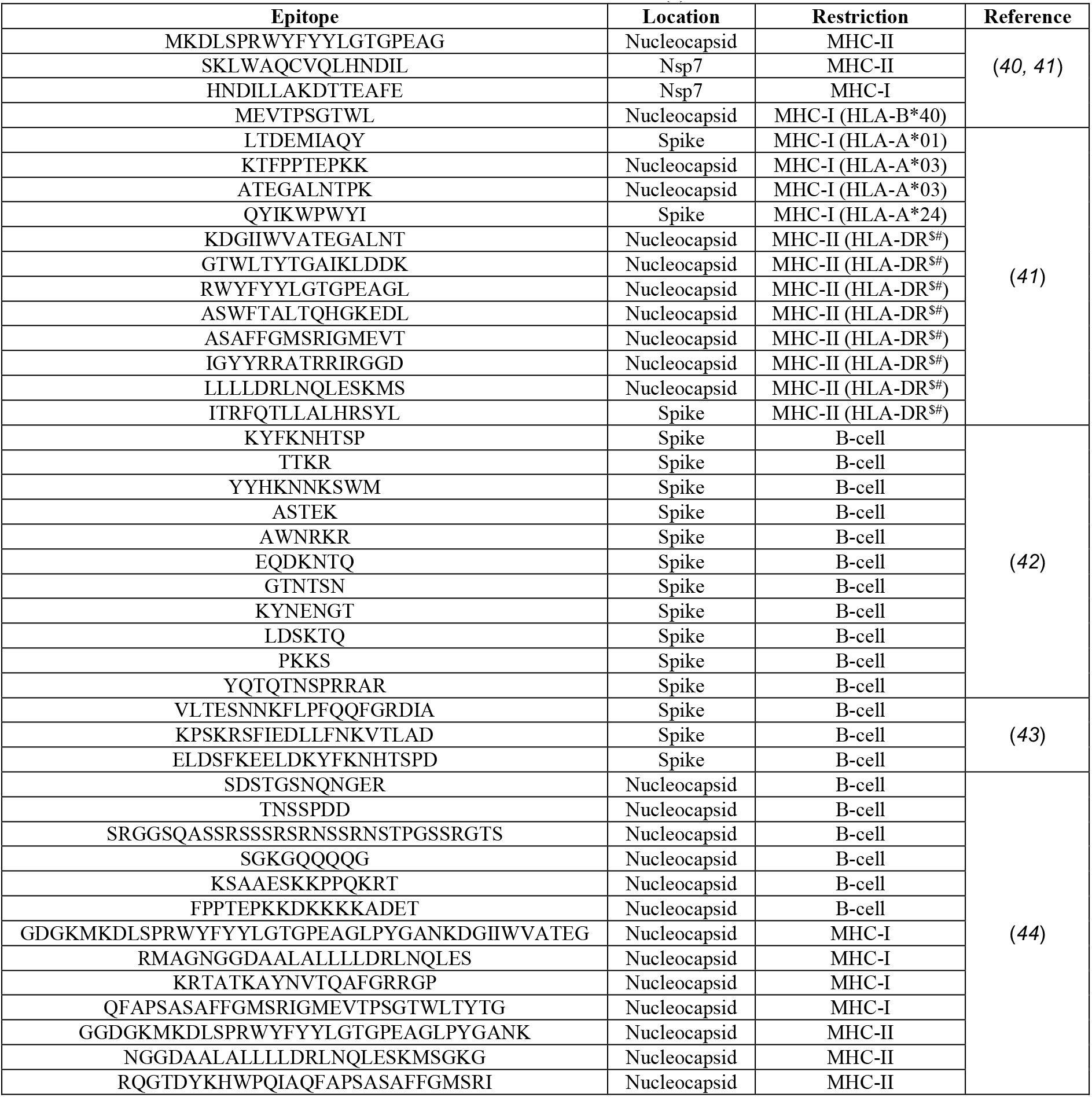
SARS-CoV-2 T-cell and B-cell epitopes in Spike and Nucleocapsid. Epitopes demonstrating cross-reactivity other human coronaviruses are denoted (^#^). In some studies, specific HLA alleles were examined for MHC-I/II. MHC-II HLA-DR allotypes included: DRB*01:01, DRB*03:01, DRB*04:01, DRB*07:01, DRB*11:01, and DRB*15:01 (^$^)

